# Hydroxychloroquine in COVID-19: An updated systematic review with meta-analysis

**DOI:** 10.1101/2020.05.14.20101774

**Authors:** Jose Chacko, Gagan Brar, Robert Premkumar

## Abstract

**Background:** Hydroxychloroquine is being administered among patients with COVID-19 infection in many healthcare systems across the world, considering its in vitro effect against the SARS-CoV-2 virus. In spite of several observational studies and a few randomized controlled trials, the effect of hydroxychloroquine on patients with COVID-19 infection remains unclear. We undertook this systematic review with meta-analysis to evaluate the efficacy and safety of hydroxychloroquine among patients with COVID-19 infection.

**Methods:** We searched PubMed, Embase, the Cochrane Library, Web of Science, medRxiv, and other relevant resources until August 1, 2020. We included randomized controlled trials and observational studies in which hydroxychloroquine was administered and compared to a control group. Data were extracted, and quality assessment of the studies was carried out. We evaluated symptomatic progression, mortality, viral clearance, evolution of changes on chest CT imaging, and adverse events. A fixed or random-effects model was used depending on outcome heterogeneity.

**Results:** We included 23 studies, including seven randomized controlled trials and 16 observational studies. Among these, 11,029 patients received hydroxychloroquine alone or in combination, while 12063 did not. Mortality was reported at different points in time. The overall mortality was not significantly different among patients who received hydroxychloroquine compared to the control group (OR: 0.94, 95% CI: 0.72–1.22; p = 0.63). Clinical worsening did not differ between patients who received hydroxychloroquine compared to those who did not (OR 0.93, 95% CI: 0.57–1.52; p = 0.77). Negative conversion, assessed by RT-PCR, did not differ significantly between the hydroxychloroquine and the control groups (OR: 0.67, CI: 0.21–2.11; p = 0.49). The evolution of changes on chest CT imaging was reported only in two studies; a more pronounced improvement was observed with the use of hydroxychloroquine compared to standard care (OR: 2.68, CI: 1.1–6.55; P = 0.03). The incidence of adverse events was significantly higher with hydroxychloroquine (OR: 5.95, CI: 2.56–13.83; p < 0.00001).

**Conclusions:** Our meta-analysis does not suggest improvement in mortality, clinical progression, or negative conversion by RT-PCR among patients with COVID-19 infection who are treated with hydroxychloroquine. There was a significantly higher incidence of adverse events with hydroxychloroquine use.

## 1. Introduction

Late last year, a novel coronavirus outbreak was identified in Wuhan, China. The SARS-CoV-2 virus spread exponentially across the globe, and the World Health Organization declared it as a pandemic in March 2020 [1]. The treatment of COVID-19 infection remains largely supportive; several treatment modalities have been proposed including the aminoquinolines, chloroquine and hydroxychloroquine [2]. Both these drugs have been extensively used to treat malaria, systemic lupus erythematosus, and rheumatoid arthritis. There has been increasing interest in the possible efficacy of these agents in COVID-19 infection, considering their anti-inflammatory and antiviral effects in vitro. The Food and Drug Administration in the US authorized emergency use of these drugs in the treatment of COVID-19 infection in March 2020, followed by extensive use across the world [3]. Hydroxychloroquine has a more potent antiviral effect and may be safer compared to chloroquine [4] and hence is more commonly used in clinical practice. Following an early report from Marseilles, France [5], which revealed more rapid viral clearance, there has been increasing interest in the efficacy of hydroxychloroquine in COVID-19 infection. However, many of these studies are limited by the lack of a control arm and are inadequate to draw definitive conclusions [6,7].

The clinical efficacy of hydroxychloroquine in patients with COVID-19 infection remains unclear despite numerous studies of limited sample size. A meta-analysis of such studies could reduce the possibility of a type II error by increasing the sample size, and may reveal any possible benefit from the intervention. Hence, we performed a systematic review and meta-analysis of available controlled studies to evaluate the safety and efficacy of hydroxychloroquine in the treatment of COVID-19 infection.

## 2. Methods

### 2.1 Search strategy and study selection

The meta-analysis was conducted in accordance with the recommendations of the Preferred Reporting Items for Systematic Reviews and Meta-Analyses (PRISMA) statement [8]. We performed a systematic search of PubMed, Embase, the Cochrane Library, Web of Science, and the medRxiv databases until August 1, 2020. Besides, we performed gray literature search using online search engines, blog search, and hand search through the table of contents of key journals. We used the keywords “COVID-19”, “SARS-CoV-2”, and “hydroxychloroquine” to search for articles. Boolean operators (AND, OR, NOT) were used as appropriate to identify relevant literature. No filters were set for the search process.

We evaluated the titles and abstracts of articles for potential study inclusion. Furthermore, the bibliography of the selected articles and previous systematic reviews were assessed for relevant articles.

### 2.2 Inclusion and exclusion criteria

Studies were considered eligible if they included patients who received hydroxychloroquine alone or in combination with other specific treatment modalities for COVID-19 infection and were compared with a control group. Both randomized controlled trials (RCTs) and observational studies with a comparator group were considered for inclusion. Data on at least one of the following outcomes had to be available for inclusion in the meta-analysis: (i) mortality, (ii) clinical progress, (iii) results of the reverse transcription-polymerase chain reaction (RT-PCR) test after the commencement of treatment, (iv) changes on computed tomography (CT) imaging of the chest, and (iv) adverse clinical events. We excluded studies in languages other than English and those with incomplete data.

### 2.3 Data extraction, assessment of study quality, and risk of bias

Data were collected independently by two authors. We collected data including the name of the first author, year of publication, study design, location of the study, the number of patients included in each group, and the dose of hydroxychloroquine administered. The outcomes evaluated included mortality at any point in time, clinical worsening, negative conversion by RT-PCR, improvement of lesions on chest CT imaging, and adverse events. The Cochrane risk of bias tool was used to evaluate RCTs [9]. The ROBINS-I tool was used to assess the risk of bias in observational studies [10]. Disagreement between investigators was resolved through discussion and consensus.

### 2.4 Statistical analysis

The outcomes studied were dichotomous; point estimates are expressed as odds ratio (OR) with the 95% confidence interval (CI). Heterogeneity of outcomes was calculated using the I^2^ statistic. An I^2^ value of 0%–40% was considered to be not important; 30% to 60% as moderate heterogeneity; 50%–90 as substantial heterogeneity and 75% to 100% as considerable heterogeneity [11]. We used a random-effects model for I^2^ ≥ 40% and a fixed-effects model for I^2^ < 40%. The meta-analysis was performed using the Mantel Hazel method as all the endpoints were dichotomous. A p-value < 0.05 was considered to be statistically significant. All analyses were performed using Review Manager 5.3 (RevMan 5.3; The Cochrane Collaboration, Oxford, UK).

## 3. Results

### 3.1 Selection of studies

We identified 1891 publications through database searching. An additional article was obtained through hand searching. We evaluated the title and abstract of 1032 articles after removing 860 duplicate publications. Of these, 938 records were excluded as they were not relevant to the meta-analysis. The full text of 94 publications was evaluated in detail; 71 of these were excluded. The excluded articles comprised of 27 letters, editorials or opinion, 20 review articles, 13 case series, six recommendations or guidelines, and five study protocols. The flow chart of study selection is depicted in Figure 1.

**Figure 1.**
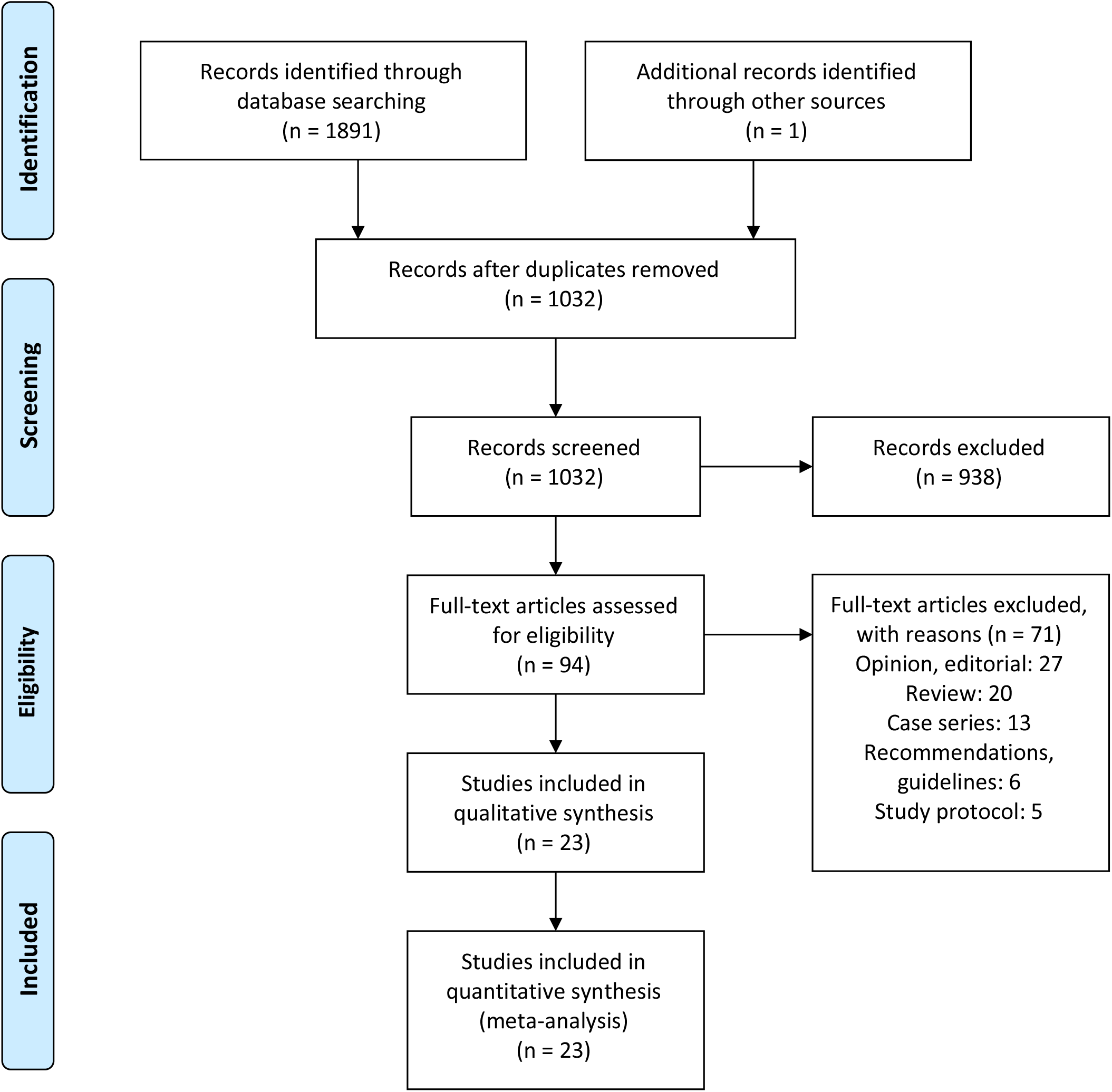
Flow diagram depicting the process of selection of the included studies.

### 3.2 Characteristics, quality, and risk of bias assessment of the included studies

The main characteristics of the studies included in the meta-analysis are presented in Table 1. Seven were RCTs [12-18] and the remaining 16 were observational studies. The included studies comprised of a total of 23,092 patients; 11,029 were in hydroxychloroquine arm, while 12,063 were in the control group.

**Table 1.** Summary of characteristics of the studies included in the meta-analysis. HCQ, hydroxychloroquine; azi, azithromycin.

| Study | Country | Design | Intervention | Control | Outcomes |
| --- | --- | --- | --- | --- | --- |
| **Cavalcanti et al. (16)** | Brazil | RCT | HCQ 400 mg twice daily, or HCQ 400 mg twice daily plus azi 500 mg daily for 7d | Standard treatment | Clinical status at day 15, hospital mortality, adverse events |
| **Chen et al. (13)** | China | RCT | HCQ 400 mg/d x 5 d | Standard treatment | RT-PCR negativity on d 7, Clinical worsening, CT changes, adverse events |
| **Horby et al. (18)** | UK | RCT | 800 mg at 0 and 6h; 400 mg at 12 h; 400 mg BD x 9d | Standard treatment | 28-d mortality |
| **Mitjà et al. (17)** | Spain | RCT | 800 mg d1, 400 mg OD X 6 d | Standard treatment | Requirement for hospitalization; adverse events |
| **Skipper et al. (15)** | US and Canada | RCT | 800 mg; 600 mg in 6–8 h, 600 mg OD X 4 d | Standard treatment | Symptom severity at 14 d, mortality, adverse events, hospital mortality |
| **Tang et al. (14)** | China | RCT | HCQ 200 mg/d x 3 d followed by 800/d | Standard treatment | RT-PCR negativity on d 28, clinical progression, adverse events |
| **Zhaowei et al. (12)** | China | RCT | HCQ 400 mg/d x 5 d | Standard treatment. Included oxygen therapy, antivirals, antibiotics, immunoglobulin, corticosteroids | Time to clinical recovery, CT changes |
| **Arshad et al. (20)** | US | Observational | HCQ 400 mg BD x 2 doses; 200 mg BD on days 2-5; azi 500 mg OD day 1; 250 OD x 4 d | 4 group: HCQ alone, azi alone, HCQ+azi, neither drug | Hospital mortality |
| **Barbosa et al. (28)** | US | Observational | HCQ 400 mg twice daily x 1–2 d; 200–400 mg/d x 3–4 d | Usual care | Mortality at 5 days, escalation of respiratory support |
| **Esper et al. (29)** | Brazil | Observational | HCQ 800 mg; 400 mg daily x 5 days. Azi 500 mg daily | Standard care | Requirement for hospitalization |
| **Gautret et al. (34)** | France | Observational | HCQ 200 mg thrice daily; azi 500 mg/d x 1d, 250 mg/d x 4 d in 6 patients | Details not available | RT-PCR negativity d 6 |
| **Geleris et al. (5)** | US | Observational | HCQ 600 mg twice daily x 1d; 400 mg daily for 4 days. Azi 500 mg x 1d; 250 mg/d x 4 d as option. Left to physician judgement. | Standard treatment | Mortality until study follow-up date. Composite of intubation or mortality on time-to-event analysis. |
| **Hraiech et al. (19)** | France | Observational | HCQ 600 mg/d; azi 500, followed by 250 mg/d | Standard treatment; no anti-viral drugs | Mortality at 6 days of ARDS onset; RT-PCR negativity d6 |
| **Ip et al. (22)** | US | Observational | HCQ dosing at physician discretion; most received 800 mg day1, 400 mg day 2–5 | Usual care; tocilizumab preferentially for ICU patients | Mortality at 30 days |
| **Kim et al. (23)** | Korea | Observational | HCQ 200 mg twice daily; antibiotics | Conservative management | RT-PCR negativity d7 |
| **Magagnoli et al. (24)** | US | Observational | HCQ alone or in combination with azi. Details of dosing not available | Standard treatment | Hospital mortality, need for mechanical ventilation |
| **Mahevas et al. (25)** | France | Observational | HCQ 600 mg/d | Standard treatment | Mortality at 7 days, need for ICU care, development of ARDS |
| **Mallat et al. (30)** | Abu Dhabi | Observational | HCQ 400 mg twice daily x 1 d, followed by | Standard treatment | RT-PCR negativity d 14 |
|  |  |  | 400 mg daily<br>x 10 days. |  |  |
| **Membrillo et al. (31)** | Spain | Observational | HCQ 800+400 mg; 400 mg daily | Standard treatment | Hospital mortality |
| **Rosenberg et al. (26)** | US | Observational | HCQ 200 mg–400 mg once or twice daily | 4 groups: HCQ alone, HCQ+azi, azi alone, and neither | Hospital mortality, cardiac arrest, abnormal ECG findings |
| **Sbidian et al. (27)** | France | Observational | HCQ 600 mg on d1; 400 mg daily for 9 days. Azi 500 mg d1; 250 mg/d x 4 days as therapeutic option | Standard care | Clinical worsening; hospital mortality |
| **Singh et al. (32)** | US | Observational | HCQ dose not specified; combined with azi in the majority of patients | Details of treatment in the control group not available |  |
| **Yu et al. (33)** | China | Observational | HCQ 200 mg twice a day x 7–10 d | Standard treatment | Hospital mortality |

The risk of bias among the included RCTs, assessed by the Cochrane risk of bias tool, is presented in Figures 2 and 3. Among the observational studies, one was considered to be at low risk, [19] eight at moderate risk, [20-27] seven at serious risk [12,28-33] and one at a critical risk of bias [5].

**Figure 2.**
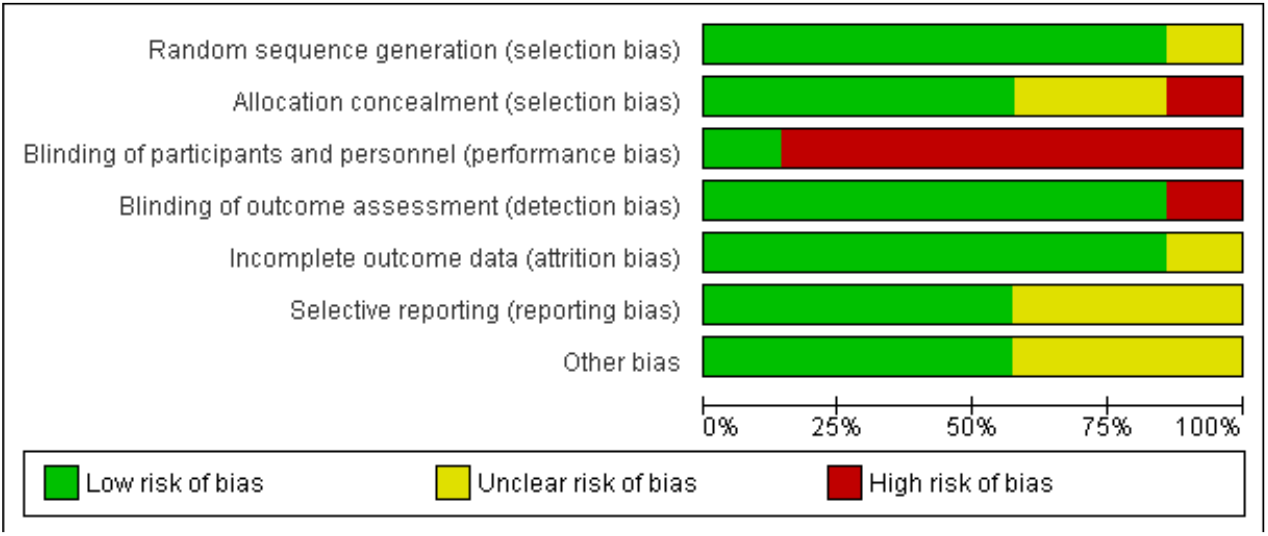
Risk-of-bias graph of randomized controlled studies using the Cochrane tool.

**Figure 3.**
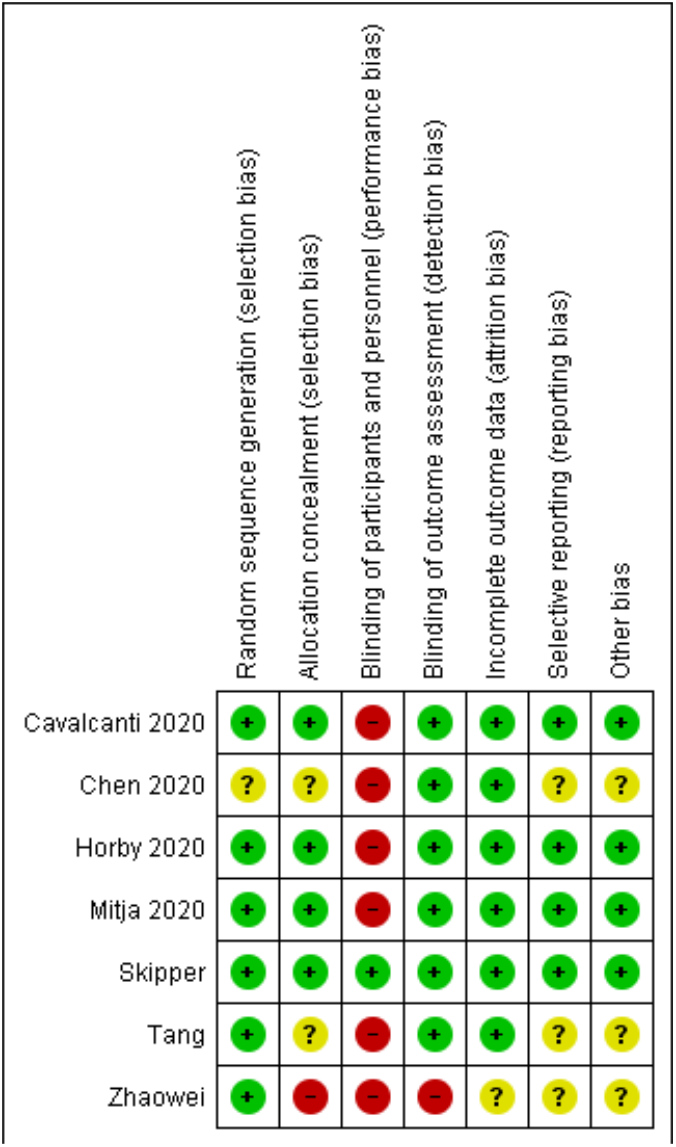
Risk-of-bias summary of randomized controlled studies using the Cochrane tool.

### 3.3 Outcomes

Fifteen studies provided data on mortality, including three RCTs and 12 observational studies Mortality was assessed at variable time points. Hospital mortality was reported by nine studies [16,20,21,24,26,28,31-33]. Two studies evaluated 28-day mortality [18,27]. Ip et al. [22] reported the 30-day and Mahevas et al. [25], the 21-day mortality. Hraiech et al. reported mortality after 6 days of onset of ARDS [19]. Skipper et al. reported two deaths in their study, one in a hospitalized and the other in a non-hospitalized patient [15]. The overall mortality was 1944 of 10,276 (18.9%) with hydroxychloroquine vs. 2432 of 11,473 (21.2%) in the control arm. Significant statistical heterogeneity was observed between studies in the evaluation of mortality (I^2^ = 86%); hence we used a random-effects model for analysis. The overall mortality was not significantly different among patients who received hydroxychloroquine compared to those who did not (OR: 0.94, 95% CI: 0.72-1.22; p = 0.63); (Figure 4). Mortality was not significantly different on pooled analysis of RCTs (OR: 1.12, 95% CI: 0.98-1.29; p = 0.10) and observational studies alone (OR: 0.9, 95% CI: 0.65-1.26; p = 0.55) (Figure 4). We performed a subgroup analysis of studies that used a daily maintenance dose of 400 mg or less of hydroxychloroquine per day compared to a dose of more than 400 mg per day. The initial loading dose was not considered in this analysis. Two studies were excluded from this analysis as the dose of hydroxychloroquine used was unclear [29,32]. No statistically significant difference in mortality was observed between a 400 mg or higher dose of hydroxychloroquine per day (OR: 0.78, 95% CI: 0.27-2.25; p = 0.65) compared to a lower dose of hydroxychloroquine (OR: 0.77, 95% CI: 0.52-1.15; p = 0.21) (Figure 5).

**Figure 4.**
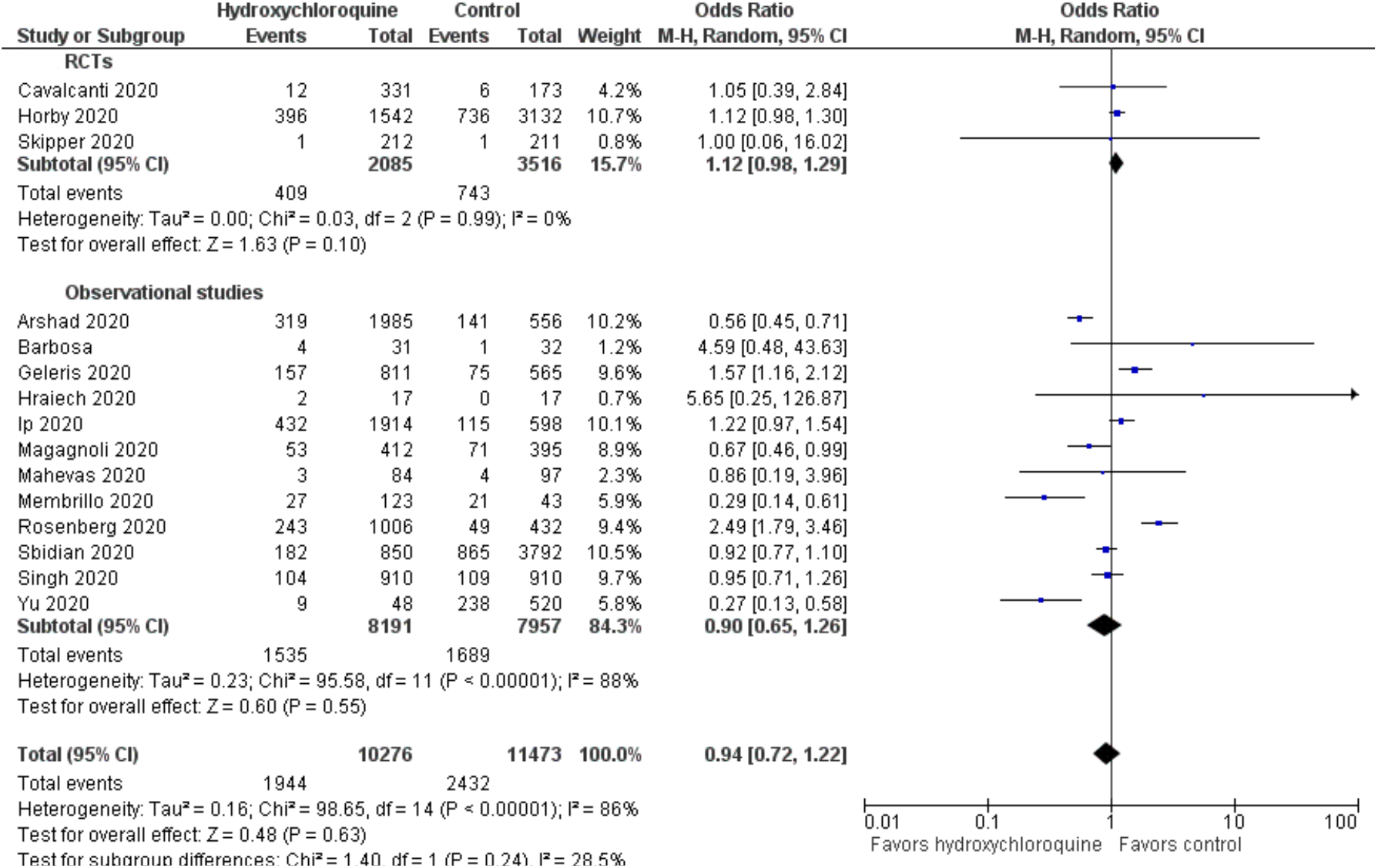
Forest plot comparing mortality between the hydroxychloroquine and the control groups. CI, confidence interval; df, degrees of freedom; M-H, Mantel–Haenszel; Random, random-effects model.

**Figure 5.**
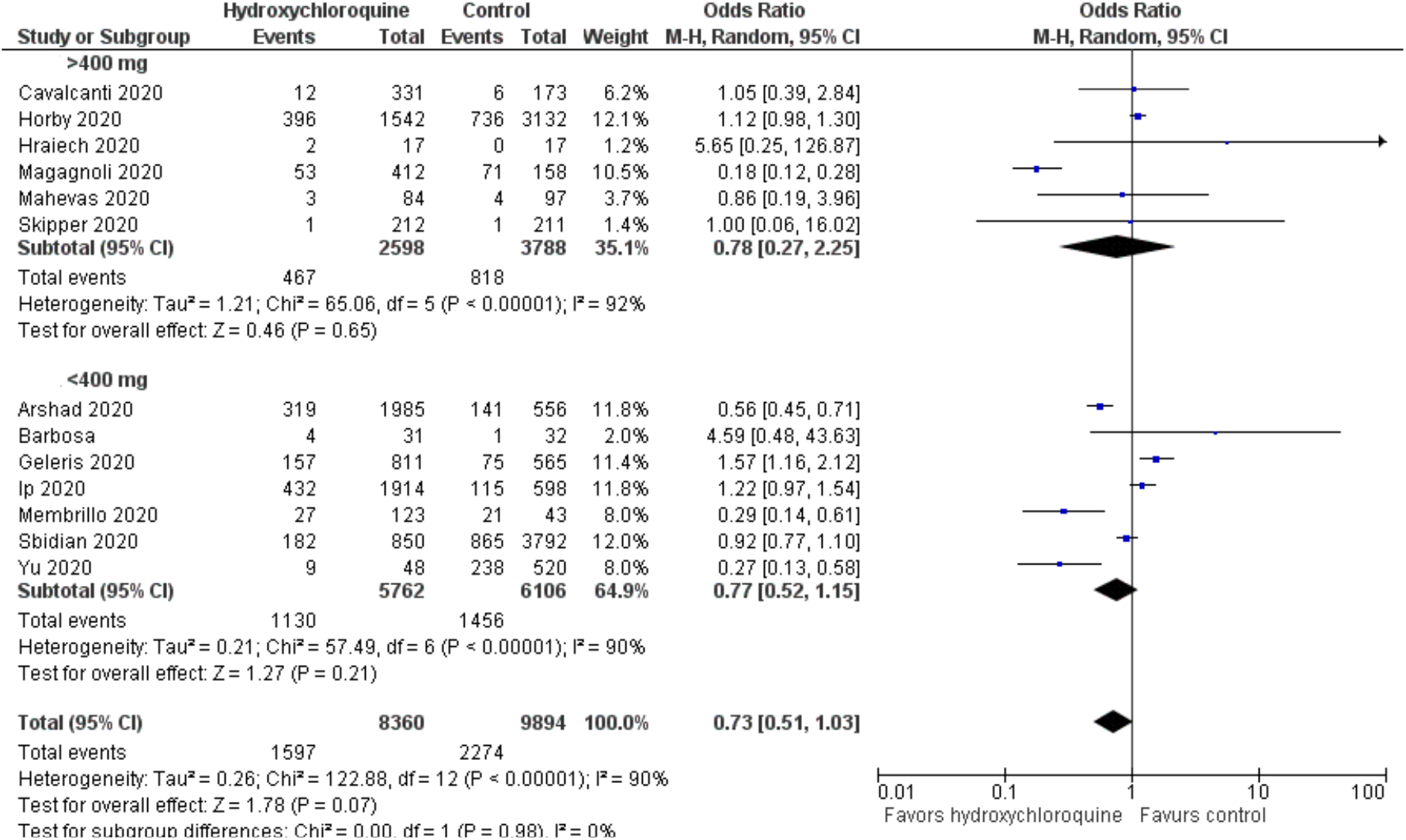
Forest plot comparing mortality with a maintenance dose of up to 400 mg per day and more than 400 mg per day of hydroxychloroquine. CI, confidence interval; df, degrees of freedom; M-H, Mantel–Haenszel; Fixed, fixed-effects model.

Clinical worsening was reported in six RCTs [12-17] and three observational studies. [27,29,32]. Clinical worsening was variously described as development of “severe” illness [12], resolution of fever and improvement in oxygen saturation [14], requirement for hospitalization [17,29] requirement for ICU transfer [27], requirement for mechanical ventilation [32], symptom severity on a 10-point visual analog scale [15], clinical status on a 7-point ordinal scale [16], and alleviation of cough with fever resolution [12]. Considerable heterogeneity was observed between studies (I^2^ = 83%); hence we used a random-effects model for analysis. Overall, there was no difference in clinical worsening between the hydroxychloroquine and the control groups (OR: 0.93, 95% CI: 0.57-1.52; P = 0.77). Pooled analysis of RCTs (OR: 0.86, 95% CI: 0.64-1.16; p = 0.32) and observational studies alone (OR: 0.99, 95% CI: 0.42-2.34; p = 0.98) did not reveal a significant difference in clinical worsening between the hydroxychloroquine and the control groups (Figure 6).

**Figure 6.**
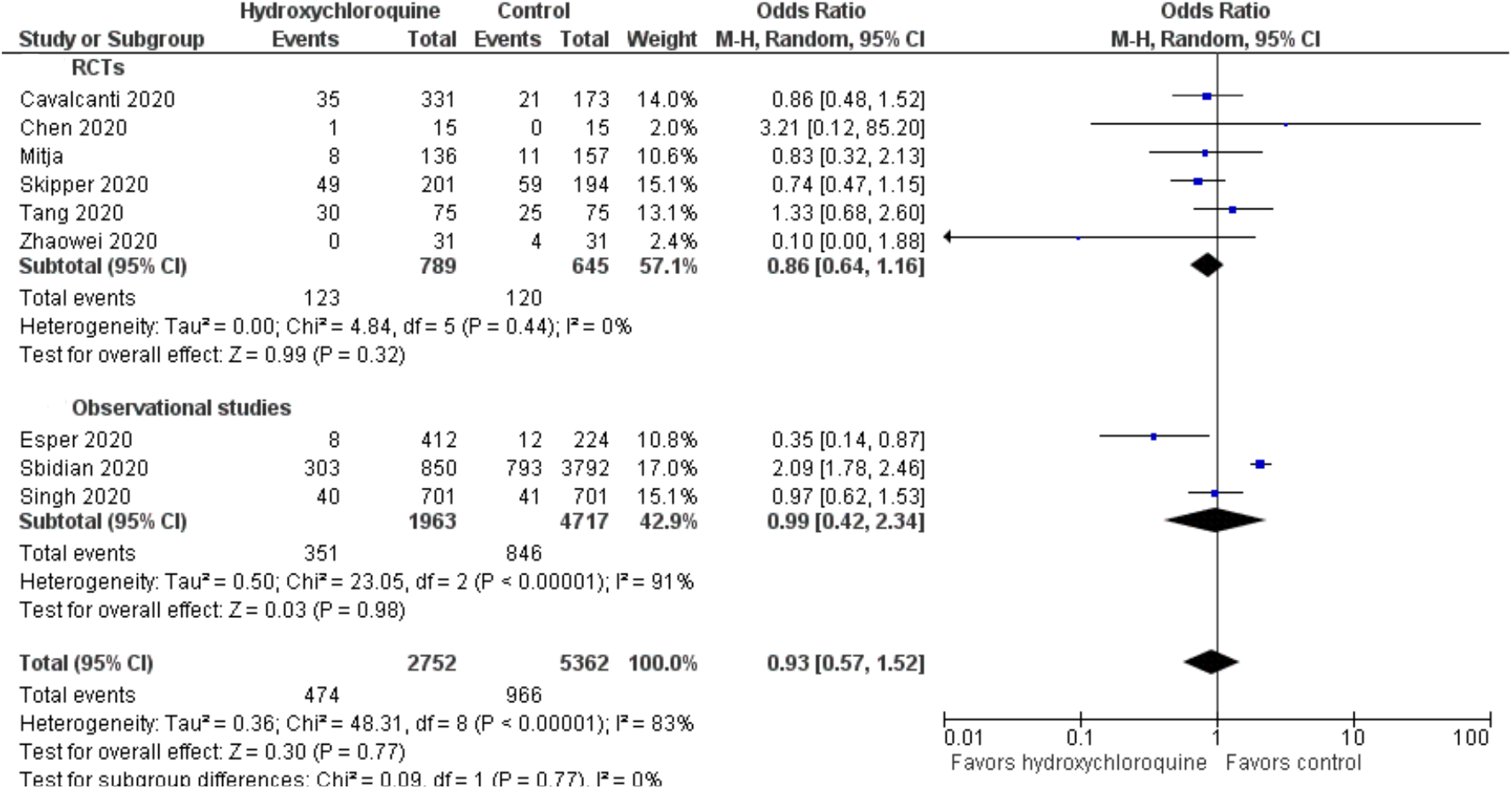
Forest plot comparing clinical worsening between the hydroxychloroquine and the control groups. CI, confidence interval; df, degrees of freedom; M-H, Mantel–Haenszel; Fixed, fixed-effects model.

Negative conversion by RT-PCR was reported at different time points in five studies. Three studies reported on negative conversion of RT-PCR by 7 days [13,23,24], while the other two studies reported at 14 [30] and 28 [14] days. There was moderate heterogeneity between studies (I^2^ = 60%); hence we used a random-effects model for the evaluation of this outcome. Negative conversion rate by RT-PCR was not significantly different between the hydroxychloroquine and control groups (OR: 0.67, CI: 0.21-2.11; p = 0.49) (Figure 7).

**Figure 7.**
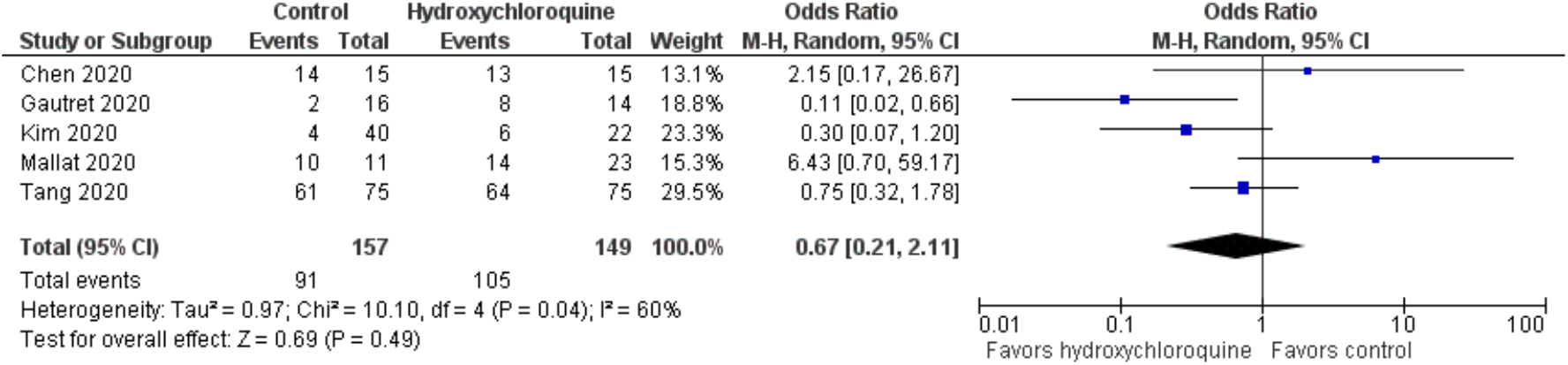
Forest plot comparing negative conversion by RT-PCR between the hydroxychloroquine and the control groups. CI, confidence interval; df, degrees of freedom; M-H, Mantel–Haenszel; Random, random-effects model.

We evaluated the improvement in changes on CT imaging of the chest, as reported in two studies [12,13]. There was no heterogeneity noted between studies (I^2^ = 0), hence we used a fixed-effect model. A more pronounced improvement on the repeat CT scan was observed with the use of hydroxychloroquine compared to standard care (OR: 2.68, CI: 1.1-6.55; P = 0.03). (Figure 8)

**Figure 8.**
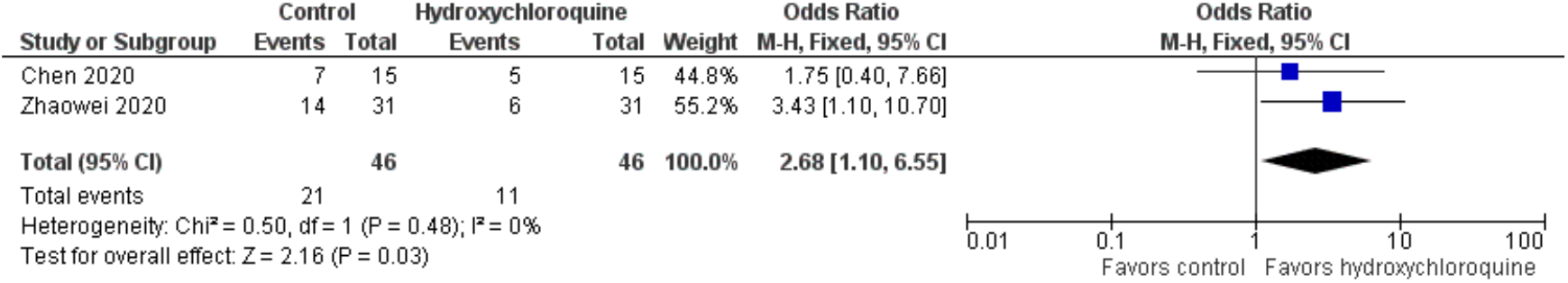
Forest plot comparing resolution of changes on CT chest between the hydroxychloroquine and the control groups. CI, confidence interval; df, degrees of freedom; M-H, Mantel–Haenszel; Fixed, fixed-effects model.

Adverse events were reported in seven studies, including five RCTs [12-15,17] and two observational studies [23,26]. There was substantial heterogeneity between studies (I^2^ = 90%); hence we used a random-effects model. Overall, adverse events were significantly more common with hydroxychloroquine compared to the control group (OR: 5.95, CI: 2.56-13.83; p = 0.001). The incidence of adverse events were higher with hydroxychloroquine on pooled analysis of RCTs (OR: 6.42, 95% CI: 1.94-21.2; p = 0.002) and observational studies (OR: 4.53, 95% CI: 0.92-22.28; p = 0.06). (Figure 9)

**Figure 9.**
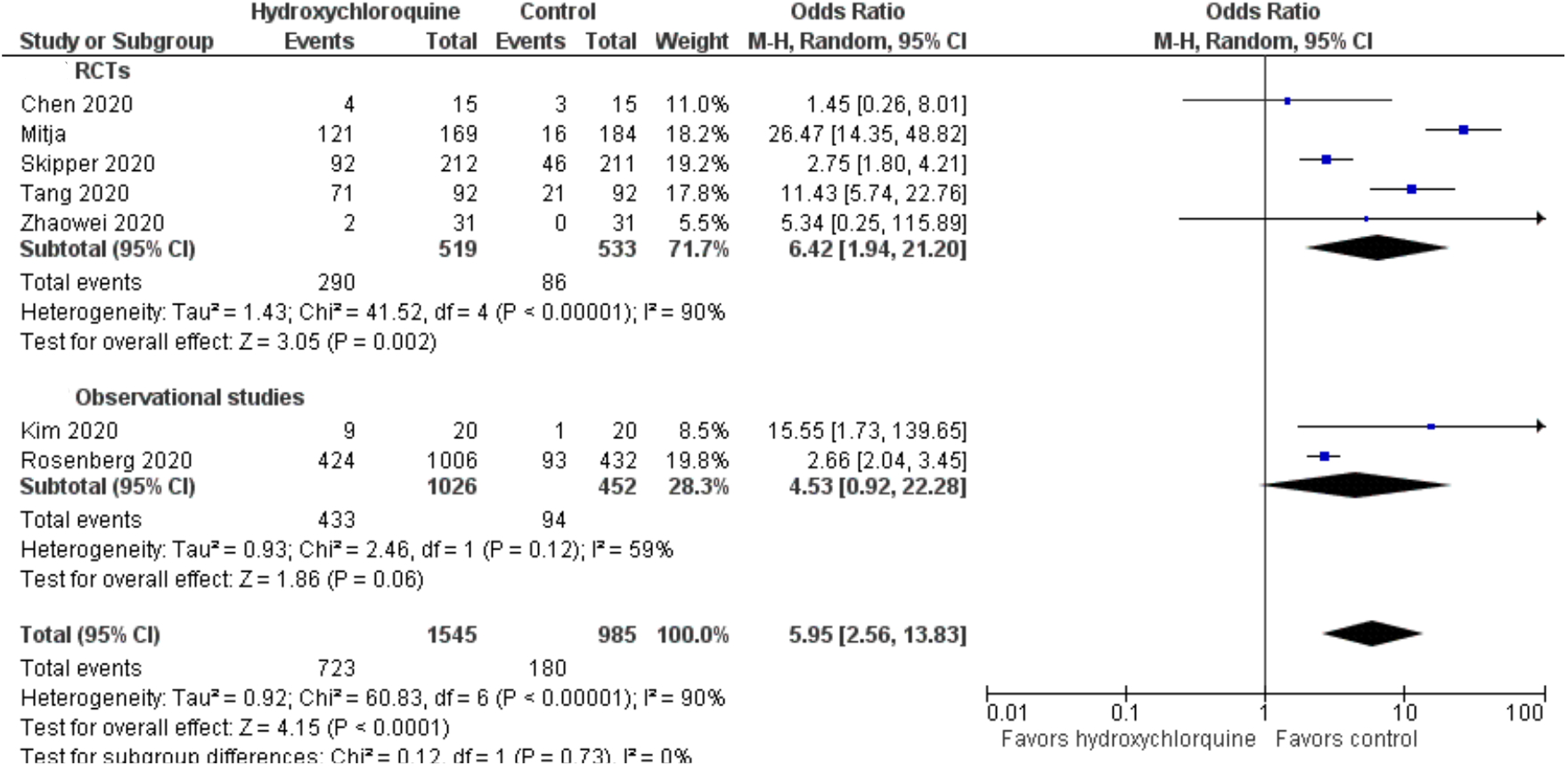
Forest plot comparing adverse events between the hydroxychloroquine and the control groups. CI, confidence interval; df, degrees of freedom; M-H, Mantel–Haenszel; Random, random-effects model.

## 4. Discussion

The synthesized evidence from our meta-analysis suggests that the use of hydroxychloroquine in patients with COVID-19 infection does not improve mortality or result in more rapid relief of symptoms. Besides, hydroxychloroquine does not appear to lead to a more rapid negative conversion by RT-PCR. Exposure to hydroxychloroquine resulted in a higher incidence of adverse events compared to patients who did not receive hydroxychloroquine.

Chloroquine and its congener, hydroxychloroquine, have revealed anti-inflammatory and antiviral effects in vitro, with the latter exhibiting more potent activity [34]. Hydroxychloroquine exerts its antiviral effect by increasing endosomal pH within the cells [35]. Besides, it inhibits glycosylation of receptors on the cell surface, which prevents binding of the SARS-CoV-2 virus to the ACE-II receptor [36]. This results in blockade of the entry pathway of the virus into the cell. Since the outbreak of the pandemic in China late last year, there has been an upsurge of interest on the clinical efficacy of hydroxychloroquine in COVID-19 infection.

In an early study from Marseilles, France, 20 patients with confirmed COVID-19 disease received hydroxychloroquine 600 mg/d; azithromycin was added based on the clinical situation. Sixteen patients from another center acted as controls. By day 3, 50% of hydroxychloroquine-treated patients tested negative for the virus by RT-PCR compared to 6.3% in the control group; by day 6, 70% among the treated group tested negative compared to 12.5% in the control group. The addition of azithromycin seemed to augment viral clearance [5]. However, the outcomes of six patients from the treatment group were not reported in this study. Clinical worsening occurred in three patients requiring ICU admission, and one patient died, while treatment was discontinued in two other patients. Three other studies evaluated the time to viral clearance by RT-PCR. These studies tested RT-PCR at different points in time; Chen et al. reported no difference in viral clearance rates on the 7^th^ day of treatment [13]. Tang et al., in their RCT, found no difference in the primary endpoint of the rate of RT-PCR negativity at 28 days with hydroxychloroquine treatment [14]. RT-PCR negativity was also comparable at days 4,7,10, and 14 days in this study. In the Mallat et al. study, RT-PCR negativity was significantly lower with hydroxychloroquine compared to the control group [30]. Our meta-analysis also revealed no effect of hydroxychloroquine on negative conversion by RT-PCR testing with the administration of hydroxychloroquine. The in vitro antiviral effect of hydroxychloroquine against the SARS-CoV-2 virus needs validation in clinical practice.

Mortality as a clinical outcome was addressed in three RCTs and 12 observational studies. Overall, there was considerable heterogeneity between studies. However, on pooled analysis of RCTs alone, the heterogeneity disappeared (I^2^ = 0). Hydroxychloroquine was administered in variable doses in different studies. In most studies, an initial loading dose was administered, followed by a maintenance dose. We compared mortality between studies that used a daily maintenance dose of 400 mg or less compared with studies that used more than 400 mg. However, the dose of hydroxychloroquine did not significantly influence mortality. It is of note that three large RCTs that evaluated mortality used a maintenance dose of more than 400 mg [15,16,18]. Three observational studies in the present meta-analysis reported a significantly lower mortality with hydroxychloroquine [20,31,33]; a maintenance dose of hydroxychloroquine 400 mg per day was used in these studies. It remains unclear whether a higher dose of hydroxychloroquine may result in adverse effects and thus offset possible clinical benefit. Besides, mortality was assessed at different time points across studies, which may have confounded the results.

Clinical worsening was assessed by six RCTs and three observational studies. The criteria used to assess the clinical status were variable. Two studies were of small sample size, with very few patients who experienced worsening of symptoms in either group [12,13]. Two large RCTs assessed clinical status around 2 weeks. Cavalcanti et al. evaluated the clinical status at 15 days on a 7-poing ordinal scale [16], while Skipper et al. assessed symptom severity on a 10-poing visual analog scale among non-hospitalized patients on day 14 of illness [15]. Neither study revealed a significant change in clinical status at 2 weeks with hydroxychloroquine administration. A large retrospective observational study from France suggested reduced requirement for ICU transfer and higher rates of 28-day discharge with hydroxychloroquine; however, this study was limited by lack of information on respiratory parameters including the requirement for oxygen therapy, invasive, or non-invasive ventilation [27].

The effect of hydroxychloroquine on chest CT imaging was assessed in two studies [12,13]. Both studies showed improved resolution of consolidation with hydroxychloroquine administration. However, these studies included a small number of patients, making the findings difficult to interpret. Besides, the clinical implications of this finding is unclear.

Adverse effects were significantly more common with hydroxychloroquine compared to the control group. These were usually mild, including headache, rash, abdominal pain, diarrhea, vomiting, and elevated liver enzymes [12,13,23]. In the study by Rosenberg et al. [26], the use of hydroxychloroquine alone or in combination with azithromycin was associated with an increase in the incidence of cardiac arrest. On an adjusted model, cardiac arrest occurred more often with the hydroxychloroquine-azithromycin combination compared to either drug alone. It is important to note that the dose of hydroxychloroquine used varied between studies, ranging from 400–1200 mg/day. A dose of more than 800 daily has been predicted to rapidly decrease viral loads compared to a dose of 400 mg daily or less based on in vitro and pharmacokinetic data; however, at higher doses, complications including prolongation of the QT-interval may occur, leading to adverse clinical outcomes [37]. The optimal dose of hydroxychloroquine for clinically important antiviral effects remains unknown and needs further research.

The combination of hydroxychloroquine with azithromycin was systematically evaluated in six studies [16,20,22,24,26,27]. In most other studies, azithromycin was administered as part of initial treatment; however data on the effect of its combination with hydroxychloroquine was not available. Hence, a meaningful assessment of the efficacy of the hydroxychloroquine-azithromycin combination compared to hydroxychloroquine alone was not feasible in the present meta-analysis. Future studies are required to address the possible benefit of this combination in COVID-19 infection.

Two previous meta-analyses have been performed to assess the efficacy of hydroxychloroquine among patients with COVID-19 infection [38,39]. However, each of these meta-analyses included only three controlled studies with a limited number of patients, and no definitive conclusions could be drawn. In contrast, the present meta-analysis included 23 controlled studies, including a much larger number of patients.

Our meta-analysis is limited by the heterogeneous nature of the studies included. We included both RCTs and observational studies, which may limit the robustness of outcome assessment. The baseline severity of illness also varied between studies. Most of the studies were of small sample size, and underpowered to evaluate the outcomes that were addressed. The endpoints, including mortality, clinical worsening, and negative conversion by RT-PCR, were reported at variable points of time, making it difficult to interpret. The dose of hydroxychloroquine used varied between studies. We were unable to assess the possible effect of using azithromycin in combination with hydroxychloroquine because data regarding the use of this combination was unavailable in most studies.

In conclusion, our meta-analysis does not support the treatment of COVID-19 infection with hydroxychloroquine. We did not observe a significant difference in mortality, clinical worsening, or negative conversion by RT-PCR with hydroxychloroquine administration. Resolution of consolidation on chest CT seems to occur more rapidly with hydroxychloroquine, although the impact on clinical outcomes remains unclear. Adverse events were significantly more with the use of hydroxychloroquine. Adequately powered RCTs are required to evaluate the possible efficacy of hydroxychloroquine in CVOID-19 infection, the optimal dosage, and possible additive effects when combined with azithromycin.

## Data Availability

The data are available with the authors

## CRediT authorship contribution statement

Jose Chacko: Conceptualization, Methodology, Data curation, Formal analysis, Investigation, Resources, Supervision, Writing - original draft, Writing - review and editing. Gagan Brar: Data curation, Investigation, Writing - original draft. Robert Premkumar: Data curation, Investigation, Writing - original draft, Writing – review and editing.

## Acknowledgement

Staff members of Majumdar Shaw Medical Center, Bangalore and Aster RV Hospital, Bangalore, for their support.

